# Assessing the Knowledge, Attitudes, and Practices (KAP) of Dengue Fever in Thailand: A Systematic Review and Meta-Analysis

**DOI:** 10.1101/2024.05.04.24306876

**Authors:** Julia Crowley, Bowen Liu, Hanan Jan

## Abstract

Dengue fever cases are on the rise in Thailand with increases in flooding events. Studies pertaining to public knowledge, attitudes, and practices (KAP) of dengue prevention have frequently been used to better understand the publics’ needs towards dengue. This study conducted a systematic review and meta-analysis of past dengue KAP studies in Thailand. Criteria were developed to determine eligibility, and reviewers independently applied these criteria to identify the 9 eligible studies for the systematic review and the 3 eligible studies for the meta-analysis. Results suggest that the majority of the studies included in the systematic review reported overall low knowledge levels towards dengue prevention in Thailand. This finding was affirmed by the meta-analysis, which concluded that the overall estimate of the proportion of participants with high knowledge of dengue prevention is only 35% (95% CI: 14%-59%). Most of the studies included in the systematic review reported positive attitudes towards dengue prevention, and this finding was also affirmed by the meta-analysis, which concluded that the pooled estimate of the proportion of positive attitudes towards dengue prevention is 61% (95% CI: 43%-77%). Lastly, the majority of studies in the systematic review reported overall poor practices towards dengue prevention. Similarly, the meta-analysis found that the pooled estimate of the proportion of good practices for preventing dengue infection is only 25% (95% CI: 22%-27%).

## Introduction

Thailand’s tropical location and topography make the Southeast Asian nation vulnerable to flood hazards [1]. Past research has examined the relationship between climate change and increases in the frequency and intensity of flooding in Thailand [2–7]. Moreover, additional research has examined the impacts of climate change-induced flood increases on several sectors within Thailand, including but not limited to agriculture [5, 8–10], housing [10–12], and public health [13].

Dengue fever is recognized as a significant public health problem that is widespread throughout Thailand [14] and exacerbated by the flooding impacts of climate change [15]. Dengue is an infectious disease that is caused by any of the four serotypes of dengue virus [16]. The virus is transmitted to humans through female *Aedes* mosquitoes and is mainly present in tropical and subtropical environments. Symptoms range from a mild fever to severe dengue hemorrhagic fever and shock syndrome [16].

Increases in dengue cases following flood events in tropical and subtropical environments were detected in previous research [17, 18]. The accumulation of stagnant water from flooding in urban areas is a major breeding ground for *Aedes* mosquitoes and the subsequent transmission of dengue [19]. Occurrences of dengue in Thailand have increased continuously over the last 60 years [20] with approximately 21,689 reported cases in 2022 [21]. The virus is now the leading cause of hospitalizations and fatalities among children, and precipitation was found to be the most influential weather variable for predicting cases of dengue in Thailand [20]. Furthermore, the high incidents of dengue place substantial economic and societal burdens on Thailand [14]. The average cost per dengue occurrence is estimated to be between 41 USDs and 261 USDs with a total annual cost estimated at 440.3 million USDs [14].

The treatment of dengue consists mainly of alleviating symptoms and avoiding complications that could potentially lead to death [22]. The development of a dengue vaccine has proven to be challenging due to the presence of four antigenically distinct dengue virus serotypes [16]. Each of these serotypes is capable of a cross-reactive and disease-enhancing antibody response against the other three serotypes. A vaccine used in dengue prevention is now available in some countries, but its reported efficacy is low in dengue naïve individuals [16]. In the United States, the dengue vaccine is only approved for use in children aged 9-16 with a previously confirmed dengue infection who are living in areas with high prevalences of the virus [23].

Such challenges in the treatment and vaccination development of dengue have called for the establishment of protocols to prevent the spread of the *Aedes* mosquito. The Ross-Macdonald model argues that effective interventions for decreasing the transmission of dengue include reducing the adult mosquito population density and the mosquito contact with humans [22]. The Centers for Disease Control and Prevention (CDC) [23] advise individuals to use insect repellent, wear loose-fitting, long-sleeved shirts and pants, and take steps to control mosquitoes inside and outside of the home. Examples of mechanisms that control mosquitoes include the use of screens on windows and doors and the regular emptying of water in items like tires and flowerpots where water can accumulate [23]. Since these measures call for specific actions at the individual and community level, several studies cite the importance of community engagement and public education in reducing the spread of dengue [24–26].

Individual vulnerability to acquiring dengue is often measured by knowledge, attitudes, and practices (KAP) surveys. Such surveys aim to measure what is known (knowledge), believed (attitude), and done (practiced) pertaining to the topic of interest [27]. KAP surveys originated in the 1950s and have become widely accepted as a research tool for health-related behaviors and health-seeking practices [27]. Given the health risks of acquiring dengue, sufficient knowledge, positive attitudes, and proper practices (KAP) are crucial for the prevention and control of the virus [28]. Furthermore, high levels of KAP can empower individuals to take part in the necessary disease control and prevention programs [29].

A myriad of studies have used KAP surveys to measure the knowledge, attitudes, and practices pertaining to dengue in different areas of the world. Many of these studies took place in Southeast Asia where the prevalence is high [30]. Past research has also provided systematic reviews and meta-analyses for the results of dengue KAP studies in individual countries. Countries in the Southeast Asia region include Malaysia [31, 32] and the Philippines [33]. However, there is a dearth of past research studies that provides systematic reviews and meta-analyses of KAP studies in Thailand. This research is crucial given the increasing cases of dengue [20] and the susceptibility of the country to increased flooding due to the effects of climate change [2–7]. This research therefore aims to examine the existing knowledge, attitudes, and practices (KAP) of dengue in Thailand by providing a systematic review and meta-analysis of past studies. The following section will describe the methods that were employed to answer the research question.

## Methods

The Preferred Reporting Items for Systematic Reviews and Meta-Analyses (PRISMA) statement [34] was used to guide this systematic review and meta-analysis. The following subsections explain the eligibility criteria, literature review, study selection, data abstraction, study appraisal, and statistical analyses.

### Eligibility Criteria

Studies were deemed eligible for inclusion in this systematic review and meta-analysis if they were observational, conducted in Thailand, and reported outcomes that included standardized scores for knowledge, attitudes, and practices in addition to specifying the proportion of participants with good knowledge, attitudes, and practices pertaining to dengue fever. Studies that did not report the results of an observational study, such as comments, case reports, reviews, and letters to the editor were excluded.

### Literature Search

Two investigators conducted separate comprehensive searches of the PubMed and MEDLINE online databases for studies published between January of 1950 and October of 2023 without language restrictions. The following terms and variants were utilized in the search strategy to identify relevant studies: “Thailand”, “KAP”, “Knowledge”, “Attitude”, “Practice”, “Dengue”, “Breakbone Fever”, “Dengue Fever.” The two investigators also independently examined the references of eligible original studies and relevant meta-analysis articles.

### Study Selection

At the initial study selection stage, two investigators independently screened the titles and the abstracts of the articles identified with the search strategy. The studies that both reviewers considered to be irrelevant to this systematic review and meta-analysis were excluded, while the remaining studies were prepared for further assessment in the next stage. During the second stage of study selection, two investigators independently examined the full texts of articles for eligibility in the meta-analysis and systematic review. Cohen’s Kappa statistic was used to evaluate the level of agreement between the two investigators at both stages of study selection. The disagreements were resolved by two additional investigators. The details of the study selection are presented in the PRISMA flow diagram in Figure 1.

**Figure 1.**
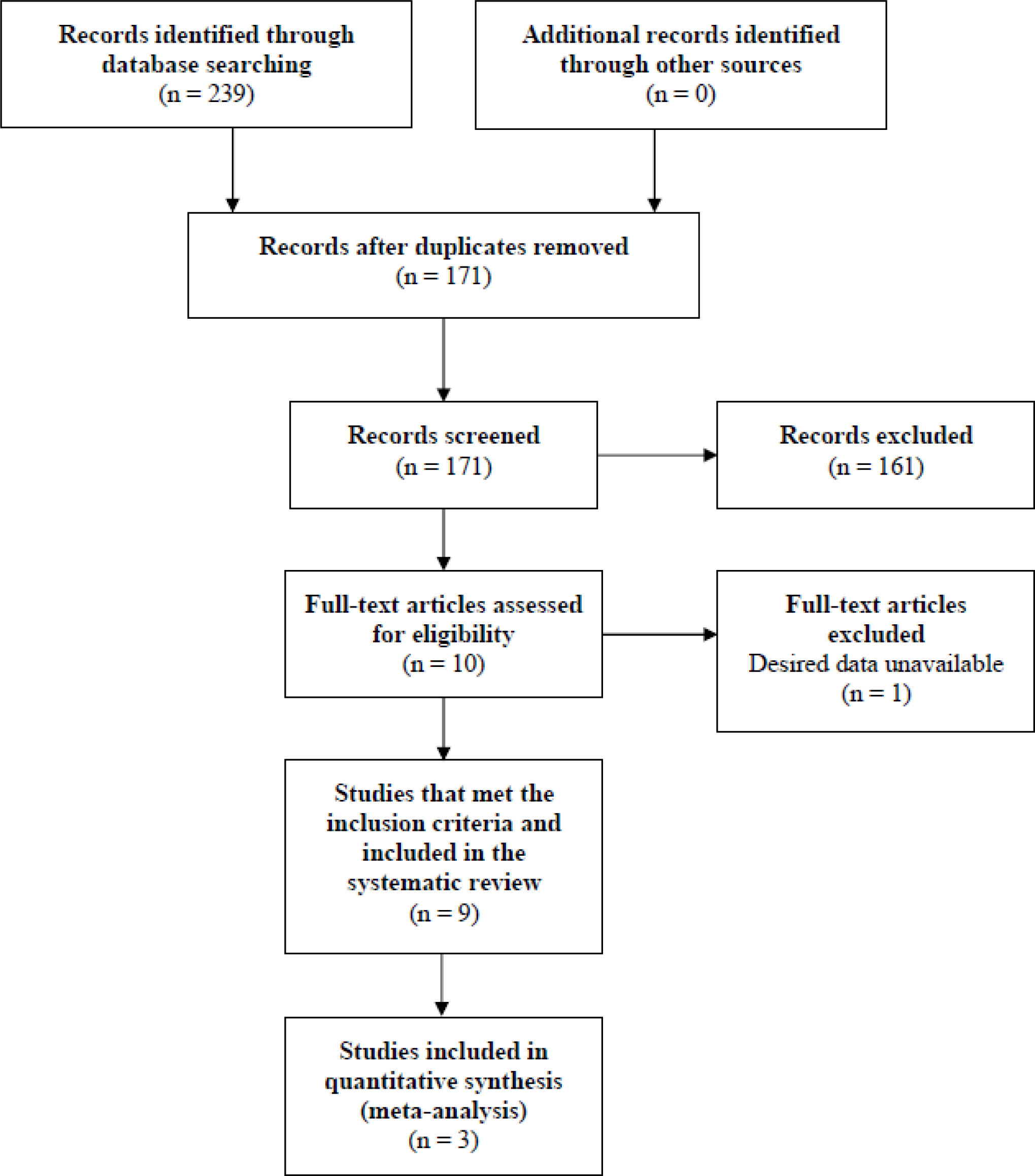
PRISMA Flow Chart

### Data Abstraction

The following study characteristics were abstracted: author(s), titles, year of publication, sample size, sampling methods, standardized knowledge scores, standardized attitude scores, standard practice scores, the proportion of study participants with good knowledge, the proportion of study participants with good attitudes, and the proportion of study participants with good practices. With a standardized data extraction form, two reviewers independently completed data abstraction and entry. The inconsistencies in data abstraction for the two reviewers were assessed by utilizing Cohen’s Kappa. Disagreements in data abstractions were resolved by two additional reviewers.

### Study Appraisal

To critically appraise the studies and evaluate the risk of bias, the studies that satisfied the inclusion criteria were examined by two independent reviewers based on the Critical Appraisal Skills Programme (CASP) Checklist [35]. The disagreements were resolved by consensus.

### Statistical Analyses

We conducted meta-analyses to quantitatively summarize the findings from different studies. The DerSimonian and Laird random-effects model was used to pool the effect sizes of included studies [36]. We utilized this model since it accounts for the between-study heterogeneity due to different study populations. For the studies that reported standardized scores of knowledge, attitude, and practice, the pooled mean standardized scores and the corresponding 95% confidence intervals (CIs) are presented. For the studies that reported the proportions of study participants with good knowledge, attitudes, and practices, the pooled proportions and the corresponding 95% CIs are presented. Subgroup analyses were performed with different criteria such as study types and sampling methods. To assess the heterogeneity in the meta-analyses, the Cochran’s Q test was utilized. In addition, the between-study heterogeneity was measured using Higgins I^2^ statistic [37]. To be consistent with previous literature, we regard I^2^ < 40% as minimal heterogeneity, 40 – 60% as moderate heterogeneity, 60 – 75% as substantial heterogeneity, and > 75% as considerable heterogeneity [38, 39].

To account for potential publication bias, we conducted Begg’s rank correlation test to help assess the presence of publication bias in the funnel plots [40]. All data analyses were conducted using the R statistical software package (Version 4.3.1, Core Team, Vienna, Austria), and a *p*-value < 0.05 was considered statistically significant. The subsequent results section presents the results of the systematic review and meta-analysis.

## Results

This section presents the results of the study selection criteria in the PRISMA flow chart of Figure 1. Subsequent subsections describe the outcomes of the systematic review and meta-analysis for the three individual components of knowledge, attitudes, and practices. Further information on the systematic review and meta-analysis is displayed in Figures 2-4 and Tables 1-2.

**Table 1:**
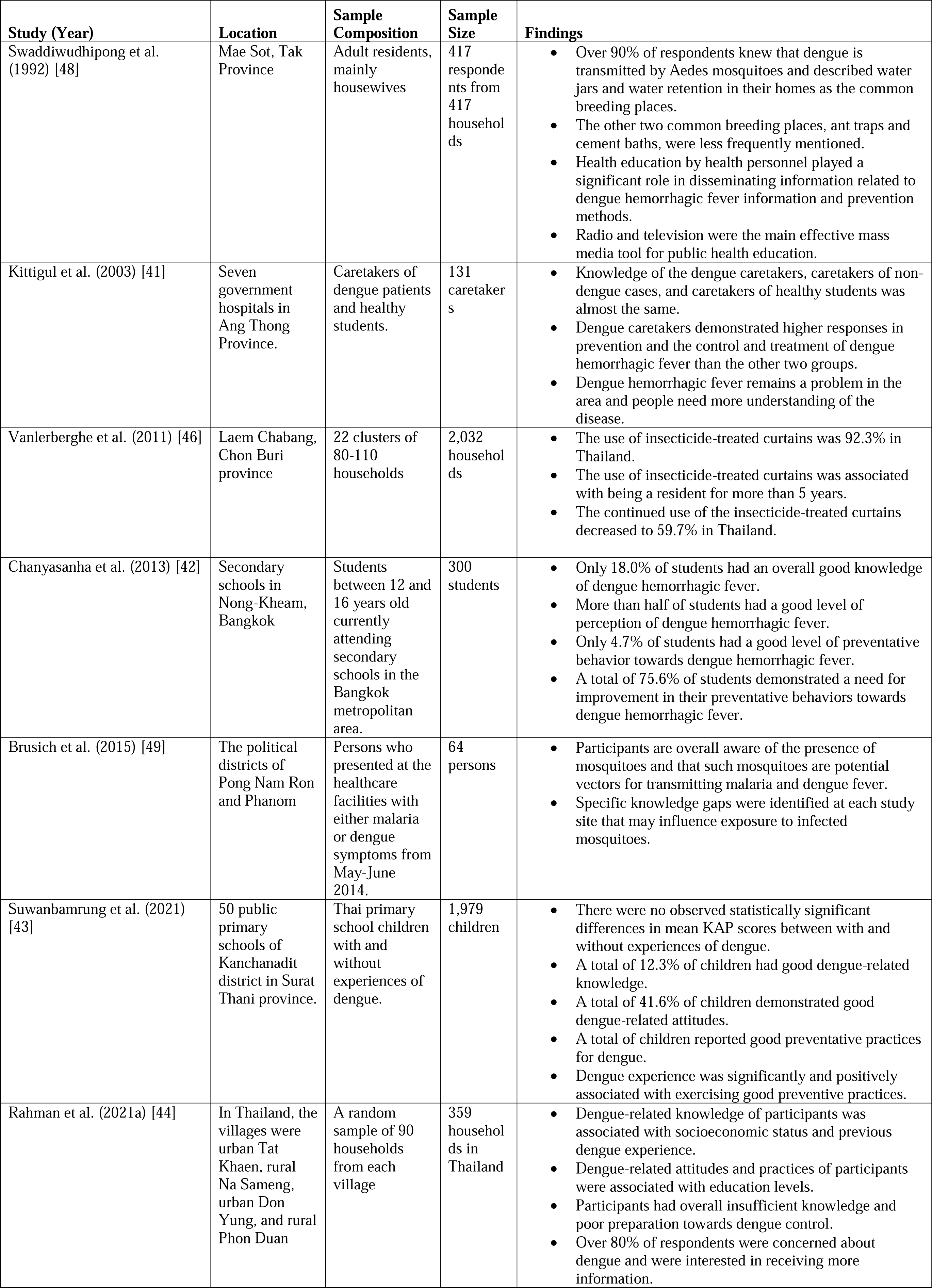
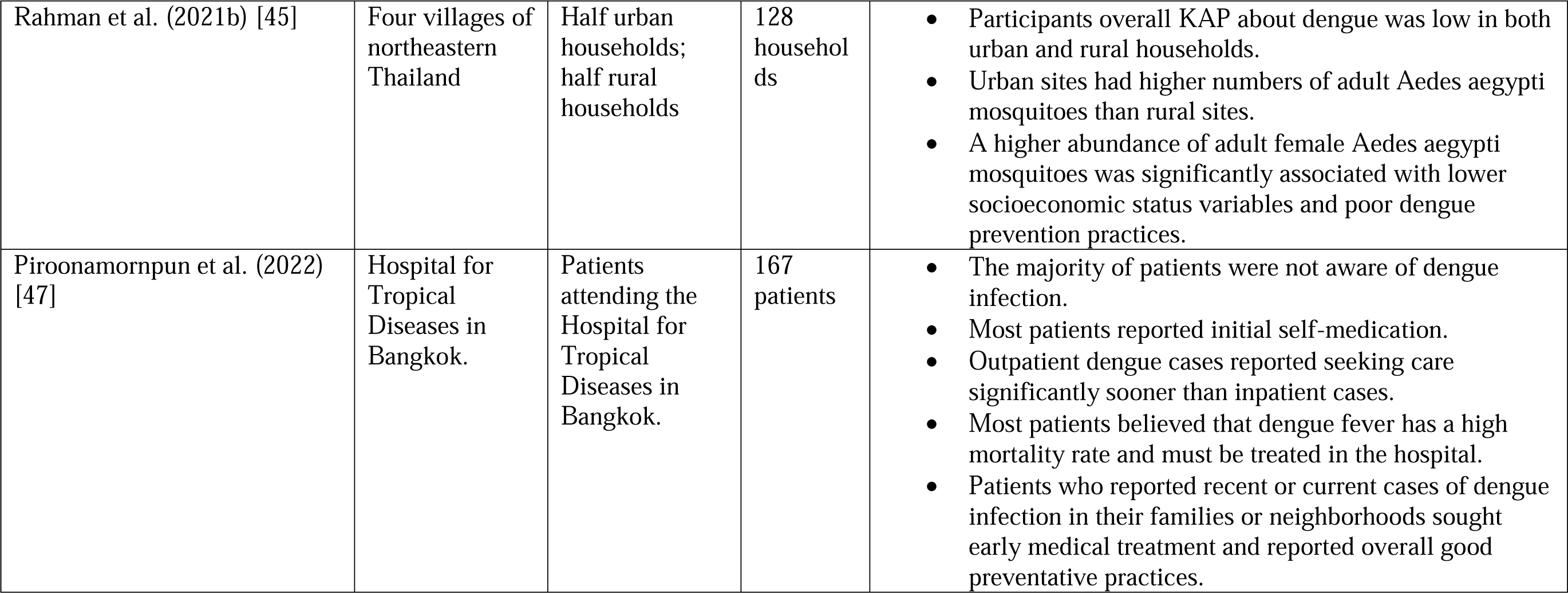
Study Characteristics.

| Study (Year) | Location | Sample Composition | Sample Size | Findings |
| --- | --- | --- | --- | --- |
| Swaddiwudhipong et al. (1992) [48] | Mae Sot, Tak Province | Adult residents, mainly housewives | 417 respondents from 417 households | Over 90% of respondents knew that dengue is transmitted by Aedes mosquitoes and described water jars and water retention in their homes as the common breeding places.<br>The other two common breeding places, ant traps and cement baths, were less frequently mentioned.<br>Health education by health personnel played a significant role in disseminating information related to dengue hemorrhagic fever information and prevention methods.<br>Radio and television were the main effective mass media tool for public health education. |
| Kittigul et al. (2003) [41] | Seven government hospitals in Ang Thong Province. | Caretakers of dengue patients and healthy students. | 131 caretakers | Knowledge of the dengue caretakers, caretakers of non-dengue cases, and caretakers of healthy students was almost the same.<br>Dengue caretakers demonstrated higher responses in prevention and the control and treatment of dengue hemorrhagic fever than the other two groups.<br>Dengue hemorrhagic fever remains a problem in the area and people need more understanding of the disease. |
| Vanlerberghe et al. (2011) [46] | Laem Chabang, Chon Buri province | 22 clusters of 80-110 households | 2,032 households | The use of insecticide-treated curtains was 92.3% in Thailand.<br>The use of insecticide-treated curtains was associated with being a resident for more than 5 years.<br>The continued use of the insecticide-treated curtains decreased to 59.7% in Thailand. |
| Chanyasanha et al. (2013) [42] | Secondary schools in Nong-Kheam, Bangkok | Students between 12 and 16 years old currently attending secondary schools in the Bangkok metropolitan area. | 300 students | Only 18.0% of students had an overall good knowledge of dengue hemorrhagic fever.<br>More than half of students had a good level of perception of dengue hemorrhagic fever.<br>Only 4.7% of students had a good level of preventative behavior towards dengue hemorrhagic fever.<br>A total of 75.6% of students demonstrated a need for improvement in their preventative behaviors towards dengue hemorrhagic fever. |
| Brusich et al. (2015) [49] | The political districts of Pong Nam Ron and Phanom | Persons who presented at the healthcare facilities with either malaria or dengue symptoms from May-June 2014. | 64 persons | Participants are overall aware of the presence of mosquitoes and that such mosquitoes are potential vectors for transmitting malaria and dengue fever.<br>Specific knowledge gaps were identified at each study site that may influence exposure to infected mosquitoes. |
| Suwanbamrung et al. (2021) [43] | 50 public primary schools of Kanchanadit district in Surat Thani province. | Thai primary school children with and without experiences of dengue. | 1,979 children | There were no observed statistically significant differences in mean KAP scores between with and without experiences of dengue.<br>A total of 12.3% of children had good dengue-related knowledge.<br>A total of 41.6% of children demonstrated good dengue-related attitudes.<br>A total of children reported good preventative practices for dengue.<br>Dengue experience was significantly and positively associated with exercising good preventive practices. |
| Rahman et al. (2021a) [44] | In Thailand, the villages were urban Tat Khaen, rural Na Sameng, urban Don Yung, and rural Phon Duan | A random sample of 90 households from each village | 359 households in Thailand | Dengue-related knowledge of participants was associated with socioeconomic status and previous dengue experience.<br>Dengue-related attitudes and practices of participants were associated with education levels.<br>Participants had overall insufficient knowledge and poor preparation towards dengue control.<br>Over 80% of respondents were concerned about dengue and were interested in receiving more information. |
| Rahman et al. (2021b) [45] | Four villages of northeastern Thailand | Half urban households; half rural households | 128 households | Participants overall KAP about dengue was low in both urban and rural households.<br>Urban sites had higher numbers of adult Aedes aegypti mosquitoes than rural sites.<br>A higher abundance of adult female Aedes aegypti mosquitoes was significantly associated with lower socioeconomic status variables and poor dengue prevention practices. |
| Piroonamornpun et al. (2022) [47] | Hospital for Tropical Diseases in Bangkok. | Patients attending the Hospital for Tropical Diseases in Bangkok. | 167 patients | The majority of patients were not aware of dengue infection.<br>Most patients reported initial self-medication.<br>Outpatient dengue cases reported seeking care significantly sooner than inpatient cases.<br>Most patients believed that dengue fever has a high mortality rate and must be treated in the hospital.<br>Patients who reported recent or current cases of dengue infection in their families or neighborhoods sought early medical treatment and reported overall good preventative practices. |

**Table 2:** Risk of Bias Assessment with the CASP Checklist.

| Study (Year) | Section A: Are the results valid? |  |  |  |  |  | Section B: What are the results? |  |  | Section C: Will the results help locally? |
| --- | --- | --- | --- | --- | --- | --- | --- | --- | --- | --- |
| CASP Questions | Was there a clear statement of the aims of the research? | Is qualitative methodology appropriate? | Was the research design appropriate to address the aims of the research? | Was the recruitment strategy appropriate to the aims of the research? | Was the data collected in a way that addressed the research issue? | Has the relationship between researcher and participants been adequately considered? | Have ethical issues been taken into consideration? | Was the data analysis sufficiently rigorous? | Is there a clear statement of findings? | How valuable is the research? |
| Swaddiwudhipong et al. (1992) [48] | Y | Y | Y | Y | Y | Y | N | N | Y | Health education by health personnel played an important role in disseminating DHF information and prevention methods. Radio and television were the main effective mass media for public health education on DHF in this area. |
| Kittigul et al. (2003) [41] | Y | Y | Y | CT | Y | Y | N | Y | Y | DHF remains a public health problem in the area and the people need more understanding of the disease. |
| Vanlerberghe et al. (2011) [46] | Y | Y | Y | Y | Y | Y | Y | Y | Y | The use of IT curtains rapidly declines over time. Continued use is mainly determined by the perceived effectiveness of the tool. This poses a real challenge if IT curtains are to be introduced in dengue control programmes. |
| Chanyasanha et al. (2013) [42] | Y | Y | Y | Y | Y | Y | Y | N | Y | The levels of knowledge, perception, and preventive behavior were low. Health education programs should be continued and intensified with emphasis on improving the knowledge of students on prevention and control practices. |
| Brusich et al. (2015) [49] | Y | Y | Y | Y | Y | Y | Y | Y | Y | Findings from this study are intended to guide future health education campaigns in these study settings to address specific community needs. |
| Suwanbamrung et al. (2021) [43] | Y | CT | Y | Y | Y | Y | Y | Y | Y | When KAP scores were categorized as good or poor levels, based on an 80% cut-off, 12.3% of all children had good dengue-related knowledge, 41.6% had good attitudes, and 25.9% reported good preventive practices. |
| Rahman et al. (2021a) [44] | Y | Y | Y | Y | Y | Y | N | Y | Y | The findings call for urgently needed integrated awareness programs to increase KAP levels regarding climate change adaptation, mitigation and dengue prevention. |
| Rahman et al. (2021b) [45] | Y | N | Y | Y | Y | CT | N | Y | Y | The low KAP regarding climate change and dengue highlights the engagement needs for vector-borne disease prevention in this region. |
| Piroonamornpun et al. (2022) [47] | Y | Y | Y | Y | Y | Y | Y | Y | Y | Health education should focus on the adult population to improve awareness of dengue symptoms and promote early treatment-seeking behavior. |

**Note: Y=Yes; N=No; CT=Can’t te**

### Knowledge

A total of 9 past studies on KAP and dengue met the inclusion criteria and were included in the systematic review. Six of the 9 included studies observed that participants had overall low knowledge pertaining to dengue [41–46] while the remaining 3 studies reported overall high participant dengue-related knowledge [47–49]. Studies commonly used awareness of the *Aedes* mosquito species as a vector for transmitting dengue to describe participants’ knowledge [43, 47–49]. Other frequently cited indictors of dengue-related knowledge include dengue symptom awareness [43, 47–49] and recognition of the time of day and season when dengue infection is most prevalent [43, 47, 48].

More specifically, studies examined associations between dengue-related knowledge and sociodemographic factors. Dengue-related knowledge was associated with participants’ socioeconomic status and 51.4% of urban participants exhibited high knowledge compared to 36.7% of their rural counterparts [44]. Other studies looked at associations between participants’ past experiences and dengue-related knowledge. The overall knowledge of dengue caretakers, caretakers of non-dengue cases, and caretakers of healthy students was almost the same between the 3 groups [41]. Dengue-related knowledge was also associated with previous dengue experiences [44], but not with the use of insecticide-treated curtains [46]. Table 1 provides details on the 9 studies and Table 2 presents the risk of bias assessment.

In total, 3 of the 9 studies reported the proportions of study participants with high knowledge of dengue [43–45]. The definition of high knowledge of dengue was consistent among the 3 studies: if a participant received a KAP knowledge score over 80%, then the participant is categorized into the group with high knowledge of dengue.

Figure 2 shows the studies that reported the proportions of participants with high knowledge of dengue. One of the studies reported the proportions separately for urban and rural areas [44]. With a random-effect meta-analysis model, the overall estimate of the proportion of participants with high knowledge of dengue is 35% (95% CI: 14%-59%). A Higgins I^2^ of 99% indicates that significant heterogeneity was observed. Publication bias was examined using Begg’s rank correlation test. The results from rank correlation tests suggested that publication bias was not significant for the proportions of knowledge (*p* = 0.17).

**Figure 2.**
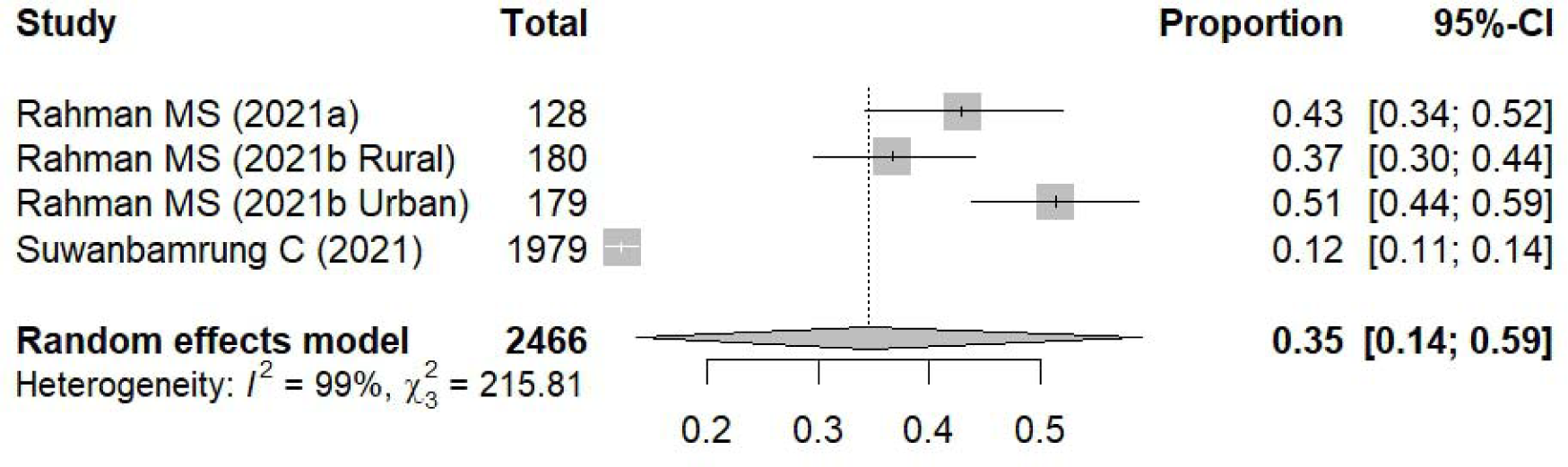
Meta-analysis of the proportion of people with good dengue knowledge in Thailand.

### Attitudes

Seven of the 9 studies included in the systematic review concluded that the majority of participants demonstrated positive attitudes towards the prevention of dengue infection through accurate perceptions of the risk of acquiring the disease [41, 42, 44–48]. The remaining 2 studies displayed negative attitudes towards dengue infection prevention through inaccurate perceptions of disease acquisition and the perceived effectiveness of preventative actions [43, 49]. Several of the studies measured attitudes by the perceived susceptibility of acquiring dengue [41–43, 47–49], the severity of the disease [41, 43, 47, 48], a desire to acquire stronger levels of dengue awareness [44, 45], and the validity of strategies to prevent infection [43–49].

Like with the knowledge component of KAP, studies also examined associations between dengue-related attitudes and sociodemographic factors. Dengue-related attitudes were associated with education levels, but the proportions of positive attitudes were almost the same between participants from rural areas (64.4%) and participants from urban areas (69.8%) [44]. Associations between participants’ past dengue experiences and dengue-related attitudes were also tested. Caretakers of dengue patients had significantly more incorrect perceptions of signs and symptoms of the disease compared to caretakers of non-dengue cases and caretakers of healthy students [41]. Finally, there was an association between the continued use of insecticide-treated curtains and their perceived effectiveness [46].

Of the 9 studies included in the systematic review, 3 reported the proportions of study participants with positive attitudes towards dengue prevention [43–45]. Three studies used the same definition for positive attitudes towards dengue prevention: If a participant received a KAP attitude score over 80%, then the participant is categorized into the group with positive attitudes towards dengue prevention.

Figure 3 demonstrates the studies that reported the proportions of participants with positive attitudes towards dengue prevention. One study reported the proportions separately for urban and rural areas [44]. The pooled estimate of the proportion of positive attitudes towards dengue prevention is 61% (95% CI: 43%-77%). A Higgin’s *I*^2^ of 97% suggested that significant heterogeneity was observed. Begg’s rank correlation test suggested that publication bias was not significant for the proportions of attitude towards dengue prevention (p = 0.17).

**Figure 3.**
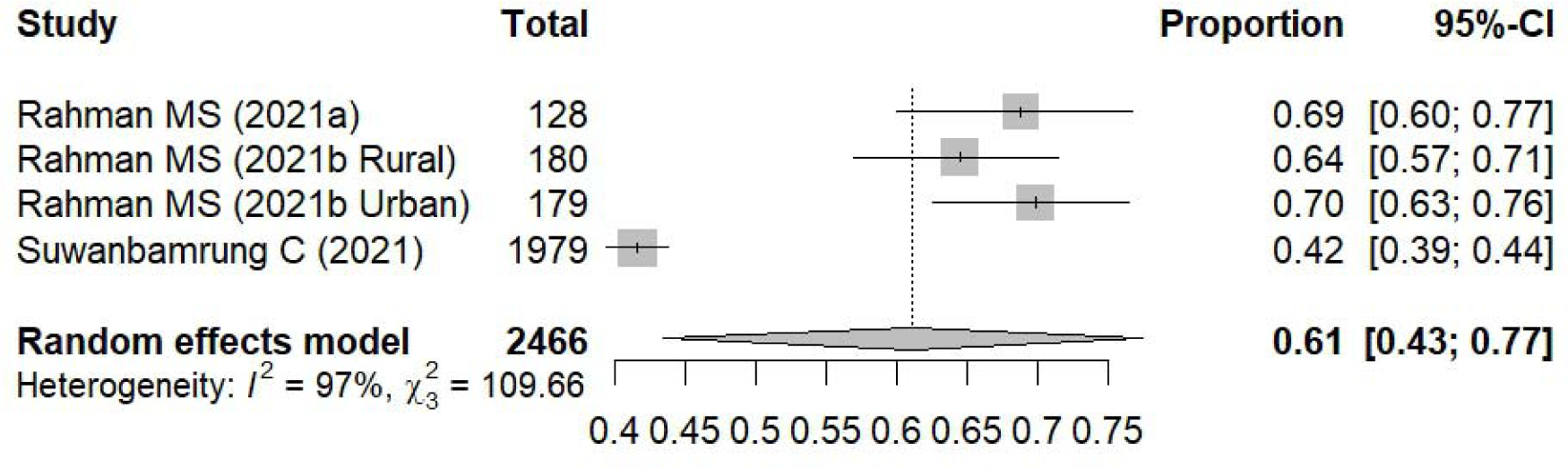
Meta-analysis of the proportion of people with good dengue attitudes in Thailand.

### Practice

Of the 9 studies included in the systematic review, only 2 of the studies displayed overall good practices for preventing dengue infection [48, 49] while the other 6 studies displayed overall poor practices [41–47]. Dengue practices in the studies commonly included the destruction of mosquito breeding sites [41–45, 47, 48], the use of pesticides [41, 44, 45], the use of mosquito nets [42, 44, 45, 48, 49], and wearing long-sleeved shirts and long pants [43–45, 49].

In addition to the knowledge and attitude components, studies looked at associations between practices and sociodemographic factors. Dengue-related practices were associated with education levels with proportions of good practices being slightly higher among urban participants (24.0%) compared to rural participants (19.4%) [44]. Past dengue experiences were also applied to practices. Caretakers of dengue patients had significantly better practices than caretakers of non-dengue cases and caretakers of healthy students [41]. Recent or current dengue infections were associated with early care-seeking and good preventive practices [47], and most participants used insecticide-treated curtains [46].

Three studies reported the proportion of participants with good practices for preventing dengue infection [43–45]. An 80% cut-off value for KAP practice scores was chosen for all three studies to determine if the study participants displayed overall good practices for preventing dengue infection.

Figure 4 presents the studies that reported the proportions of participants with good practices for preventing dengue infection. The pooled estimate of the proportion of good practices for preventing dengue infection is 25% (95% CI: 22%-27%). Higgin’s *I*^2^ of 26% suggested that heterogeneity was not significant. Begg’s rank correlation test suggested that publication bias was not significant for the proportions of good practices for preventing dengue infection (p = 1.00). The following section will provide a detailed discussion of the results.

**Figure 4.**
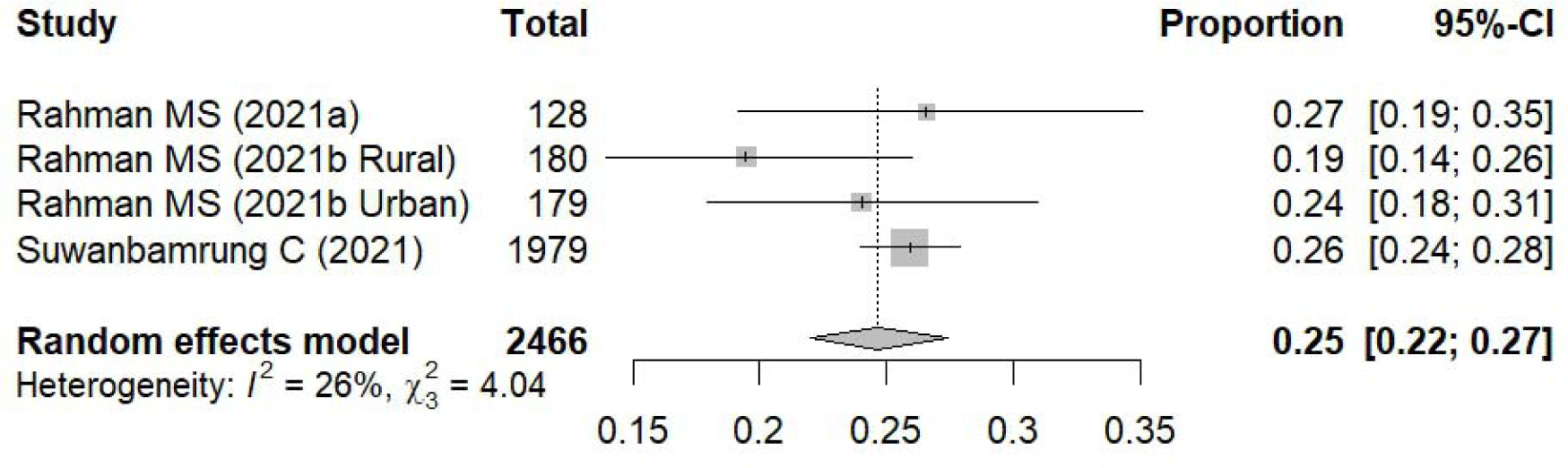
Meta-analysis of the proportion of people with good dengue practices in Thailand.

## Discussion

The overall low knowledge and poor practice levels pertaining to dengue prevention that were detected in the systematic review and meta-analysis are alarming given the previously cited rates of increasing dengue cases throughout Thailand [20, 21] and the subsequent burdens the virus imposes on the Thai economy [14]. However, the overall positive attitudes towards dengue prevention that were also observed in the results are reassuring. This finding suggests that there is a high likelihood that participants will be open to dengue education and prevention programs given their overall high-risk perceptions of the virus.

The results of this study displayed both similar and conflicting information when compared to similar studies conducted in other countries in Southeast Asia. For example, the previously cited meta-analysis on dengue KAP in the Philippines presented similar findings pertaining to knowledge with 31.1% of the studies displaying high knowledge [33], which is close to the figure of 35.0% for this study.

However, positive attitude levels were lower at 50.1% compared to 61.0% in this study while good practice levels were higher at 35.3% compared to 25.0% in this study [33]. However, greater contrasts were observed for knowledge and practices in the results of the other previously cited meta-analysis on dengue KAP in Malaysia with 51.0% of the studies exhibiting high knowledge and 45.0% displaying good practices [31]. On the other hand, attitude levels were similar with the meta-analysis in Malaysia finding that 56.0% of the studies depicted positive attitudes [31]. Furthermore, the previously cited systematic review of dengue KAP in Malaysia found significant associations between knowledge and dengue experiences, which correlates with the systematic review of this study [32].

An additional study provided a scoping review on dengue KAP studies that included geographical representation in Southeast Asia, South Asia, the Caribbean, and South America [50]. In contrast to this study and the meta-analyses in the Philippines [33] and Malaysia [31], they detected that the majority of respondents had high knowledge levels towards dengue fever [50]. However, the scoping review focused on comparing dengue KAP in dengue epidemic areas to those of controlled dengue areas and did not include a systematic review and/or a meta-analysis [50].

Other systematic reviews and meta-analyses on KAP and different types of mosquito-borne diseases generated comparable results to this study. A systematic review on the global primary literature of risk perceptions, attitudes, and knowledge of chikungunya found overall higher knowledge of the disease in areas that had previous outbreaks of chikungunya [51]. They also concluded that the majority of the populations studied did not understand chikungunya and were therefore less likely to protect themselves from mosquito bites [51]. On a similar note, a systematic review on malaria KAP in South Asia found that a general knowledge of the disease was mostly lacking among the public and healthcare professionals [52].

One limitation of this study is that the survey questions pertaining to knowledge, attitudes, and practices are not standardized across each of the studies that were included in the systematic review and meta-analysis. It is recommended that the Ministry of Public Health of Thailand work with local public health professionals within their agency, the private sector, and academic institutions to develop a standardized guide of the most crucial survey questions that should be included to measure knowledge, attitudes, and practices towards dengue prevention. This would allow for more opportunities to derive meaningful results across studies through systematic reviews and meta-analyses.

The overall positive attitudes towards dengue prevention that were observed by this study suggest that there is a strong willingness for the public to take the necessary steps to mitigate their risks towards exposure to dengue. However, the overall low knowledge levels and poor practices towards dengue prevention indicate that more public health campaigns are needed to educate the public on the risk of dengue and mitigation techniques.

## Conclusion

To conclude, the majority of the studies included in the systematic review reported overall low knowledge levels towards dengue prevention in Thailand. This finding was affirmed by the meta-analysis, which concluded that the overall estimate of the proportion of participants with high knowledge of dengue prevention is only 35% (95% CI: 14%-59%). Most of the studies included in the systematic review reported positive attitudes towards dengue prevention, and this finding was also affirmed by the meta-analysis, which concluded that the pooled estimate of the proportion of positive attitudes towards dengue prevention is 61% (95% CI: 43%-77%). Lastly, the majority of studies in the systematic review reported overall poor practices towards dengue prevention. Similarly, the meta-analysis found that the pooled estimate of the proportion of good practices for preventing dengue infection is only 25% (95% CI: 22%-27%).

In addition to the previously discussed limitation on the lack of standardization in KAP survey questions across studies, another limitation of this study is that it only examined dengue KAP studies in Thailand. Further research should conduct similar systematic reviews and meta-analyses on dengue KAP in other regions of the world and compare the results to this study.

## Statements and Declarations

The authors declare no conflicts of interests.

## Data Availability

Data sharing does not apply to this research since no datasets were generated.

